# Development and fit for purpose validation of a quantitative LC-MS/MS method for heparan sulfate in cerebrospinal fluid as a biomarker for mucopolysaccharidosis type IIIA

**DOI:** 10.64898/2026.03.27.26348847

**Authors:** Cory Bystrom, Kevin Douglass, Manju Gupta

**Author notes:** Corresponding author: Manju Gupta, PhD, Ultragenyx Pharmaceutical Inc., 60 Leveroni Court, Novato, CA 94949, USA.

## Abstract

**Background:** Mucopolysaccharidosis type IIIA (MPS IIIA; Sanfilippo syndrome) is a fatal neurodegenerative lysosomal storage disorder caused by impaired degradation of heparan sulfate (HS). Despite rapid advances in gene and enzyme therapies, there remains a critical need for an analytically validated, quantitative biomarker that accurately reflects central nervous system (CNS) substrate burden. Such biomarker would be a valuable tool in assessing disease progression and monitoring therapeutic efficacy.

**Objective:** This study describes the method development, fit for purpose validation, and preliminary clinical application of a quantitative liquid chromatography−mass spectrometry (LC−MS/MS) assay for the HS-derived disaccharide N-sulfoglucosamine–glucuronic acid (GlcNS−GlcUA) in human cerebrospinal fluid (CSF), a critical biomarker for diagnosis, disease monitoring, and regulatory evaluation of emerging MPS IIIA therapies.

**Methods:** A structurally defined GlcNS−GlcUA reference standard and its [^13^C_6_]-labeled internal standard were used in a derivatization and detection workflow employing 1-phenyl-3-methyl-5-pyrazolone labeling, and LC-MS/MS.

**Results:** The method exhibited acceptable linearity across 0.005−0.500 nmol/mL (r ≥0.9976), with intra- and inter-assay imprecision ≤3.5%CV and accuracy within 95%−110% of nominal concentrations. No matrix or hemolysis interference or carryover was observed, and the analyte remained stable during freeze-thaw storage conditions. Application of the method to 12 CSF samples from patients with MPS IIIA demonstrated quantifiable GlcNS−GlcUA levels ranging from 0.0054 to 0.106 nmol/mL, confirming suitability for clinical and regulatory use. Comparison of the MPS IIIA sample results between the development laboratory and the contract research organization laboratory support robust inter-lab assay transfer.

**Conclusions:** This validated LC–MS/MS method establishes a regulatory-grade quantitative assay for measurement of CSF HS in MPS IIIA. Its high analytical sensitivity and reproducibility enable reliable assessment of CNS substrate reduction and pharmacodynamic response, supporting biomarker-driven therapeutic development and accelerated approval pathways for neuronopathic mucopolysaccharidoses.

## 1. Introduction

Mucopolysaccharidosis type III (MPS III; Sanfilippo syndrome) is a neurodegenerative lysosomal storage disorder caused by deficient degradation of heparan sulfate (HS), resulting in progressive neurocognitive decline and premature death [1]. Clinical symptoms typically emerge between 2 and 6 years of age, followed by rapid neurologic deterioration during childhood and death in the second or third decade of life [1]. Although no disease-modifying therapy is currently approved, multiple investigational approaches, including enzyme replacement, substrate reduction, hematopoietic stem cell transplantation, and gene therapy, are advancing through mid-to late-stage clinical trials [2].

Diagnosis of MPS III typically begins with detection of elevated urinary glycosaminoglycans, such as HS, followed by molecular genetic testing and confirmatory lysosomal enzyme assays (MPS IIIA: Nsulfoglucosamine sulfohydrolase; MPS IIIB: alpha-N-acetylglucosaminidase; MPS IIIC: heparan-α-glucosaminide N-acetyltransferase; and MPS IIID: N-acetylglucosamine-6-sulfatase) [3]. Conventional dye-binding screens (eg, 1,9-dimethylmethylene blue) lack specificity and often yield false results, leading to diagnostic delays [4]. Recent liquid chromatography−tandem mass spectrometry (LC−MS/MS)−based quantification of HS in cerebrospinal fluid (CSF) has demonstrated that CSF HS concentrations are markedly elevated and decline following CNS-directed therapies in patients with MPS IIIA, suggesting that CSF provides a direct and disease-relevant matrix for assessing CNS substrate burden [5].

Recognition of the biomarker potential of CSF HS increased following the 2024 Reagan-Udall United States Food and Drug Administration (U.S. FDA) workshop, which identified CSF HS as a CNS-proximal surrogate endpoint reasonably likely to predict clinical benefit [6]. Unlike plasma or urinary HS, which primarily reflect peripheral disease activity, CSF HS mirrors central pathological processes.

Saville and colleagues identified the urinary HS-derived disaccharide HN-UA-1S, compositionally consisting of one glucosamine, one iduronic acid, and one sulfate modification, that was selectively elevated in MPS IIIA [7, 8]. Although not fully structurally resolved at that time, an LC-MS/MS assay using ΔDi-4S chondroitin sulfate as a single-point calibrator enabled semi-quantitative measurement of this fragment [7]. Subsequent studies confirmed its specificity as a secondary biomarker complementing enzyme and genetic testing [7, 8]. Collectively, the work by Saville and colleagues established the foundation for integrating HS-derived disaccharides as clinically meaningful biomarkers for MPS IIIA.

Building upon this foundation, Gholap and colleagues recently defined the complete molecular structure of HN-UA-1S and synthesized N-sulfoglucosamine–glucuronic acid (GlcNS−GlcUA) along with an isotopically labeled internal standard [9]. These advances enabled us to develop a fully characterized, quantitative LC-MS/MS assay that was further validated at an external laboratory. In the present work, we report the methodology development and fit for purpose validation of a quantitative assay to measure GlcNS−GlcUA in CSF to advance biomarker-driven drug development in MPS IIIA.

## 2. Materials and Methods

### 2.1 Reagents and Materials

All reagents and solvents used were of analytical or LC-MS/MS grade quality. Acetonitrile, methanol, high-performance liquid chromatography (HPLC) grade water, and formic acid (Optima LC-MS/MS grade) were obtained from Thermo Fisher Scientific (Waltham, MA, USA). The HS disaccharide reference standard (GlcNS−GlcUA) and its isotopically labeled internal standard ([^13^C_6_]-GlcNS−GlcUA) were synthesized and supplied by Enfanos, Inc. (Seattle, WA, USA). Chloroform, glacial acetic acid, and ammonium hydroxide (28%–30% w/w) were obtained from Thermo Fisher Scientific. The derivatization reagent 3-methyl-1-phenyl-5-pyrazolone (PMP) was purchased from TCI Chemicals (Portland, OR, USA). Pooled human CSF matrix confirmed negative for metabolic or infectious disease markers was sourced from Innovative Research (Novi, MI, USA). All reagents were used as received without further purification. CSF samples were obtained from patients with MPS IIIA who participated in a phase 1/2 clinical study (ClinicalTrials.gov, NCT02716246). The Nationwide Children’s Hospital Institutional Review Board roster approved the study protocol and all study procedures. Written, informed consent was obtained from participants before study procedures. All samples were collected following anonymized before analysis and used in accordance with ethical guidelines and regulatory requirements.

### 2.2 Preparation of Derivatization Reagent

Derivatization of HS disaccharides was achieved using PMP to enhance chromatographic retention and ionization efficiency. A 250-mM PMP derivatization solution in methanol was diluted in equal volume with 0.4 M ammonia in methanol before addition to CSF.

### 2.3 Cerebrospinal Fluid Sample Derivatization and Preparation

Samples of CSF, calibration standards, and quality controls (QC) were thawed at room temperature and gently mixed before processing. Aliquots of 100 µL of each sample, double blank, negative controls, and QC samples, were transferred into individual wells of a 96-deep-well polypropylene plate. Each well received 25 µL of the working internal standard ([^13^C_6_]-GlcNS−GlcUA). Next, 100 µL of freshly prepared derivatization reagent was added to each sample, and the plate was vortex-mixed to ensure homogeneity. The wells were then sealed and heated at 70±5°C for 90±5 minutes using a controlled heat block. Following incubation, samples were cooled to room temperature and neutralized with 200 µL of 0.6 M acetic acid. Excess PMP reagent was removed by three sequential liquid–liquid extractions using ∼300 µL chloroform per extraction. After the final extraction, the aqueous supernatant was transferred to a 0.2-µm filter unit and centrifuged at 3,000×g for 1 minute. The clarified filtrate was transferred to autosampler vials or a 96-well analytical plate, sealed with snap caps, and placed in the autosampler for LC-MS/MS analysis.

### 2.4 Liquid Chromatography–Mass Spectrometry Analysis

Quantitative analysis of PMP-derivatized GlcNS−GlcUA and its isotopically labeled internal standard ([^13^C_6_]-GlcNS−GlcUA) was performed using a UHPLC system (Thermo Fisher Scientific) coupled to a QTRAP^®^ 6500+ triple quadrupole mass spectrometer (SCIEX, Framingham, MA, USA) operated in negative electrospray ionization mode. Data acquisition and processing were carried out using SciexOS™ software.

Separation was achieved on a pentafluorophenyl (PFP) column (2.1×100 mm; 3 µm particle size) maintained at 30°C. Mobile phase A consisted of water with 0.1% formic acid, and mobile phase B was acetonitrile with 0.1% formic acid. The flow rate was 0.30 mL/min, with a total run time of 7.5 minutes. The gradient was programmed as follows: 0.0–3.2 min, 20–22.5% B; 3.2–4.2 min, 90% B; 4.2–5.2 min, hold at 90% B; and 5.2–7.5 min, hold at 20% B for re-equilibration. The injection volume was 10 µL. A divert valve was used to direct the flow to waste during the initial and final segments of each.

The mass spectrometer was operated in multiple reaction monitoring (MRM) mode under unit resolution for both Q1 and Q3. The monitored transitions were m/z 764 → 331 for quantitation and m/z 764 → 269 for qualification of GlcNS−GlcUA, and m/z 770 → 337 (quantitation) and m/z 770 → 274 (qualification) for the isotopically labeled internal standard. Ion ratio agreement was evaluated during development, but single quantifiers were used for final validation. Each transition was acquired with a dwell time of 100 milliseconds. Ion source parameters were optimized as follows: curtain gas 30 psi, ion spray voltage −4500 V, source temperature 300°C, GS1 30 psi, GS2 30 psi, and collision gas set to medium. Compound-dependent parameters were declustering potential −80 V, entrance potential −10 V, collision energy −45 V, and collision cell exit potential −25 V.

Each analytical sequence began with system suitability injections followed by calibrators, QCs, and unknown CSF samples. Additional QCs were interspersed throughout the sequence to monitor analytical stability. Calibration was established as linear regression with 1/x^2^ weighting and concentrations of GlcNS−GlcUA were reported in nmol/mL.

### 2.5 Method Validation

The initial method was developed and characterized at Ultragenyx Pharmaceutical Inc. (Novato, CA, USA). The method was transferred and subsequently validated under Good Laboratory Practice (GLP) conditions at Veloxity Labs (Peoria, IL, USA). All validation activities at Veloxity Labs were performed in accordance with U.S. FDA 21 CFR Part 58 and the FDA Guideline on Bioanalytical Method Validation for Biomarkers for Industry [10]. The study was executed according to a predefined validation plan, with all raw data, calculations, and summary results verified by the bioanalytical laboratory.

Selected performance characteristics were calibration curve linearity and range, intra- and inter-assay accuracy and imprecision, selectivity, matrix and hemolysis effects, carryover, and stability. Calibration standards ranging from 0.005 to 0.500 nmol/mL were prepared in water after demonstration of calibration equivalence between control CSF and water. Water was selected as the calibration matrix to avoid risk of bias from pooled normal CSF containing low but frequently measurable amounts of GlcNS−GlcUA. Quality control samples at 0.0050 (lower limit of quantitation [LLOQ]), 0.015 (low), 0.150 (mid), and 0.375 (high) nmol/mL were prepared in pooled authentic human CSF. Selectivity was assessed using at least six independent CSF lots. Hemolysis effects were evaluated by analyzing low- and high-QC samples spiked with 2% hemolyzed whole blood. Carryover was monitored by injecting blank matrix samples immediately following the high (ULOQ) calibrator. Freeze–thaw stability was assessed over three complete freeze–thaw cycles, each separated by ≥12 hours at ≤−20°C and ≥1 hour at ambient temperature. To ensure the applicability of the method in a real-time setting, twelve individual sources/lots of MPS IIIA patient samples were analyzed in single replicate at the development and validating laboratories.

## 3. Results and Discussion

The LC-MS/MS assay for GlcNS−GlcUA in CSF was successfully developed by Ultragenyx Pharmaceutical Inc. and fully validated under GLP compliance at Veloxity Labs. The method demonstrated high selectivity, sensitivity, and reproducibility across the analytical range of 0.005– 0.500 nmol/mL, enabling accurate quantification of HS at clinically relevant concentrations observed in patients with MPS IIIA.

### 3.1 Calibration Curve and Sensitivity

For GlcNS−GlcUA, calibration standards demonstrated a consistent and proportional response over the analytical range of 0.005–0.500 nmol/mL. Weighted (1/x^2^) linear regression model yielded a correlation coefficient (r) of 0.9998, indicating excellent linearity across the full dynamic range (**Figure 1**), with back-calculated concentration accuracies across all calibration levels (**Table 1**) ranging from 98.2 % to 104.5 % with imprecision (%CV) values ≤6.1. The LLOQ was established at 0.005 nmol/mL.

**Table 1.**
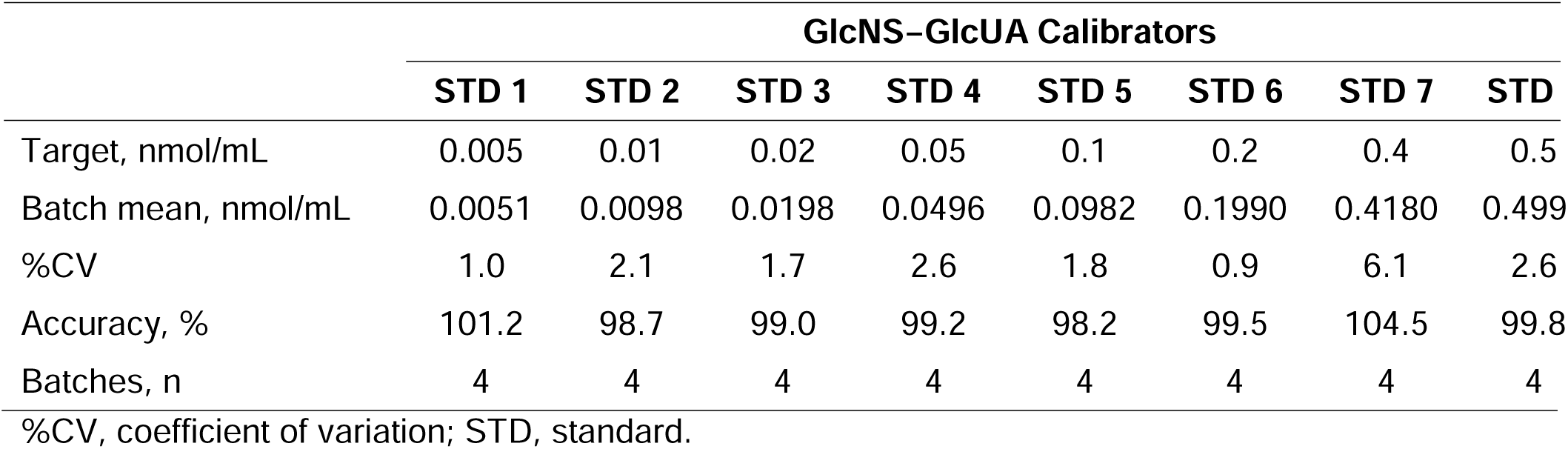
GlcNS−GlcUA Calibrator Performance.

**Figure 1.**
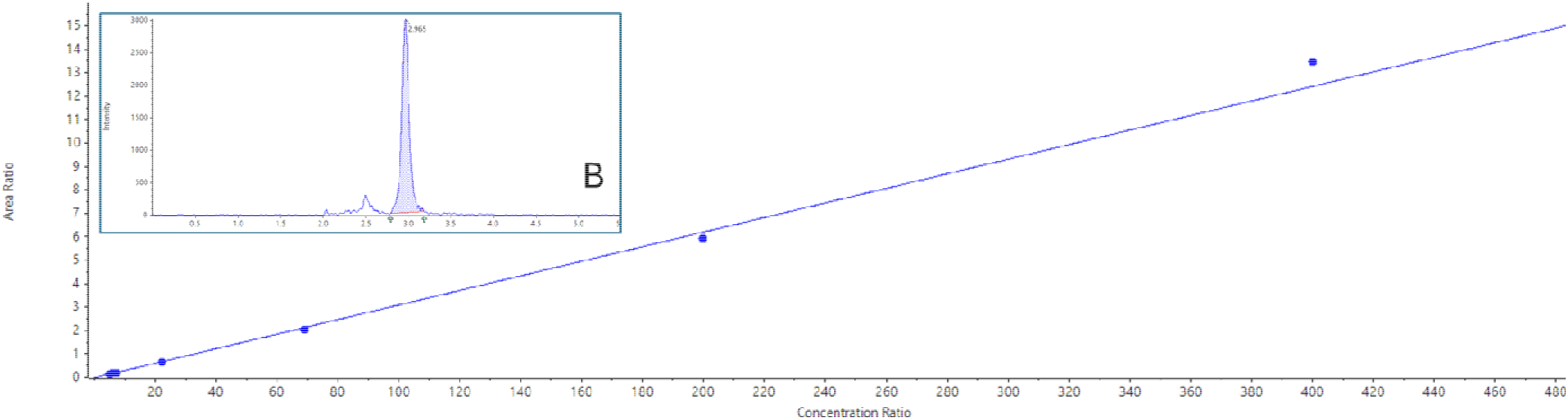
Example calibration (A) with low calibrator chromatogram (B).

### 3.2 Imprecision and Accuracy

Good precision and accuracy of the LC-MS/MS assay for GlcNS−GlcUA in human CSF was observed across multiple validation batches using QC samples. The intra-assay imprecision (%CV) ranged from 0.9% to 2.6%, while inter-assay imprecision ranged from 1.9% to 3.5% across all QC levels (**Table 2**). Mean intra- and inter-assay accuracies were within 93.3%–114.0% and 95.2%–110.2% of nominal values, respectively.

**Table 2.**
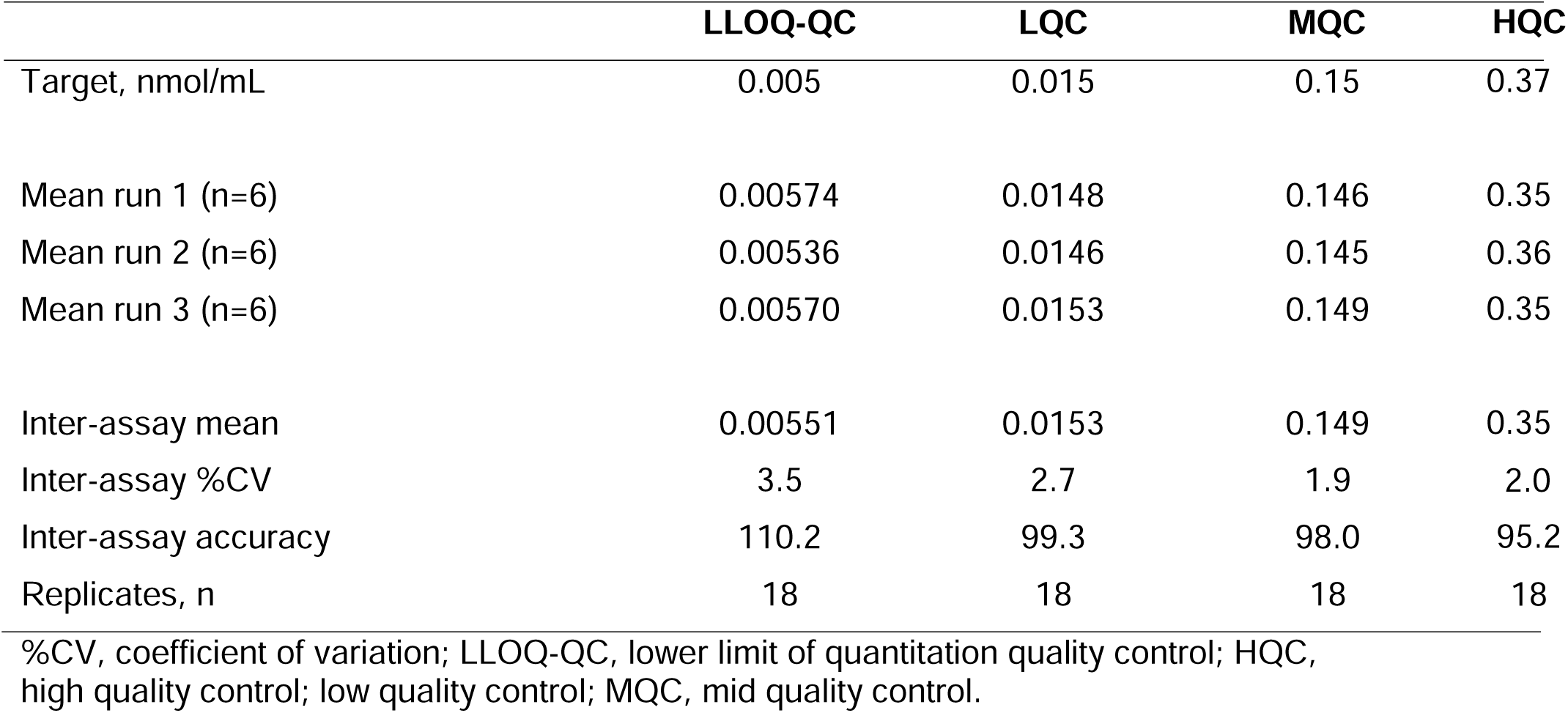
GlcNS−GlcUA Imprecision and Accuracy.

### 3.3 Selectivity, Matrix Effect, Carryover, and Ion Ratios

#### 3.3.1 Selectivity

Selectivity of the LC-MS/MS assay for GlcNS−GlcUA and its isotopically labeled internal standard ([^13^C_6_]-GlcNS−GlcUA) was confirmed in six independent lots of blank human CSF. No significant interference from endogenous matrix components was observed at the analyte or internal-standard retention times. Analyte responses in normal CSF ranged from 2.6% to 18.0 % of the LLOQ (≤20% criterion), while internal-standard responses were negligible (0% of the LLOQ signal). Representative chromatograms of matrix blanks demonstrated clean baselines with no co-eluting peaks. During development, ion ratio measurements and retention times for the synthetic calibrator agreed with quality controls and unknowns within ±10% and <0.05 minutes, respectively.

#### 3.3.2 Hemolysis effect

No hemolysis effect was observed when 2% (v/v) whole blood was added to blank matrix before spiking with low- and high-QC levels. Mean accuracy was 108% for low QC (LQC) and 99.2% for high QC (HQC), with imprecision of 1.3%CV and 1.6%CV, respectively. Although signal suppression (<50 % of normal response) was noted, quantitative accuracy remained unaffected.

#### 3.3.3 Carryover

Acceptably low carryover was demonstrated, with a mean analyte signal of 3.3%–6.4 % of the LLOQ observed in blanks after injection of the high calibrator. No mean analyte signal was observed for the internal standard.

### 3.4 Stability

#### 3.4.1 Freeze−thaw stability

Stability of GlcNS−GlcUA in CSF for at least three freeze–thaw cycles was observed with accuracy across LQC and HQC levels ranging from 99.7% to 108.7%, and imprecision (%CV) ≤1.4%.

#### 3.4.2 Benchtop and long-term stability

Twenty-hour benchtop storage was demonstrated at LQC and HQC levels. Accuracy ranged from 101.9% to 108.0% with imprecision ≤1.7%. Twenty-eight-day long term frozen storage (−20°C) was similarly determined. Accuracy ranged from 104.7% (LQC) to 99.5% (HQC), with imprecision ≤2.3%.

### 3.5 Application to MPS IIIA Patient Samples and Transferability

To demonstrate the applicability of the validated LC-MS/MS assay under real-world conditions, twelve CSF samples from patients with MPS IIIA were analyzed for GlcNS−GlcUA levels using the method in both the development and validating laboratories. Calculated concentrations of GlcNS−GlcUA in MPS IIIA CSF samples ranged from 0.0054 to 0.106 nmol/mL, demonstrating the assay’s capability to quantify the biomarker across a broad dynamic range encompassing both low- and high-burden disease states. Representative chromatograms from a patient sample are provided in **Figure 2**. Transferability between laboratories was assessed comparing results collected in the research and GLP-compliant laboratories. Linear regression revealed a slope near unity and a negligible intercept (m=1.048; 95% CI: 0.92 to 1.17; b=0.0003; 95% CI: −0.007 to 0.007; r2=0.973), whereas a difference Bland-Altman plot demonstrated one sample at the 95% agreement limit (**Figure 3**). Although limited, these findings confirm that the method performs reliably in authentic clinical matrix and is well suited for translational, clinical, and regulatory applications, including biomarker qualification and therapeutic monitoring in MPS IIIA clinical studies.

**Figure 2.**
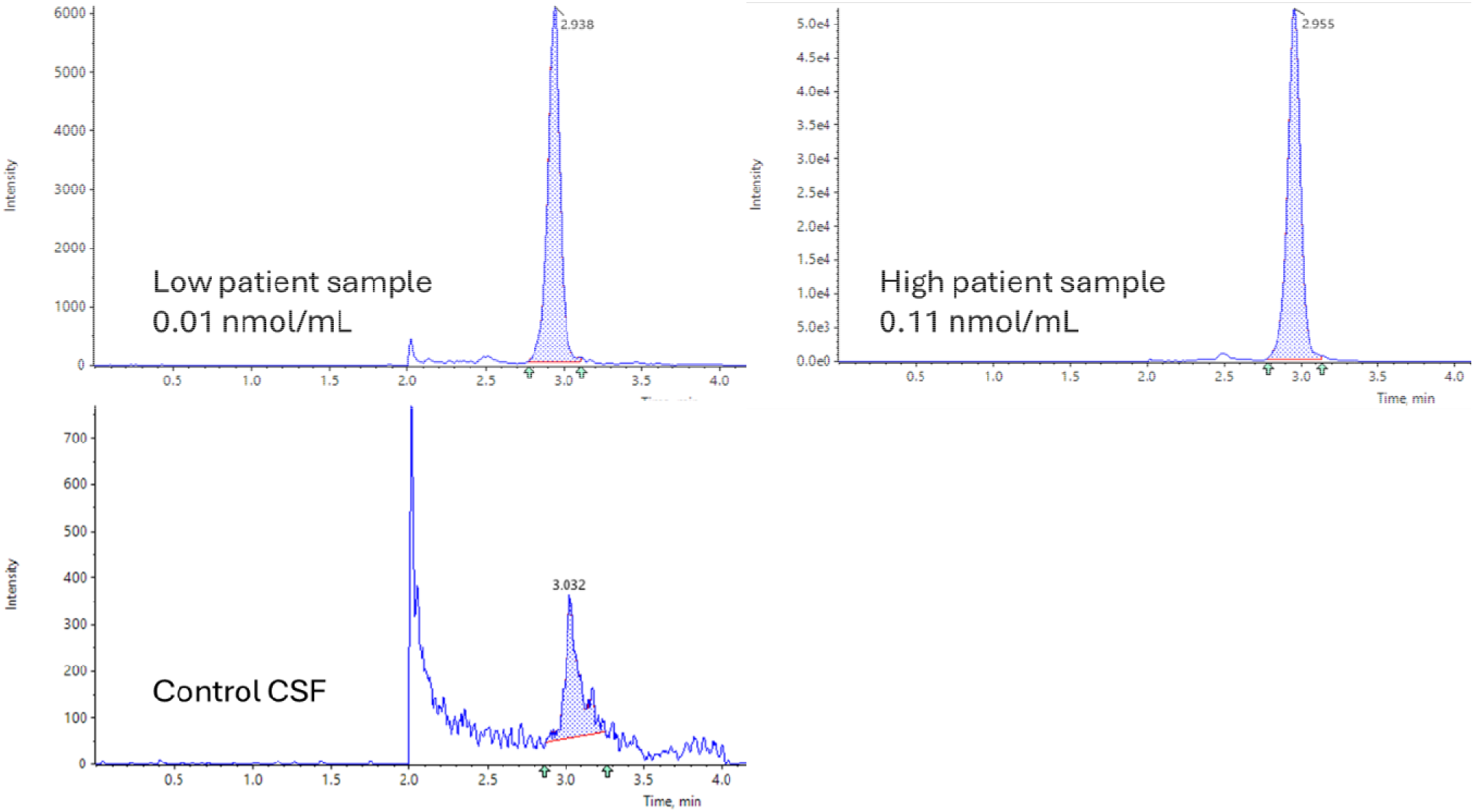
Representative chromatograms from a low, high, and control MPS IIIA patient sample.

**Figure 3.**
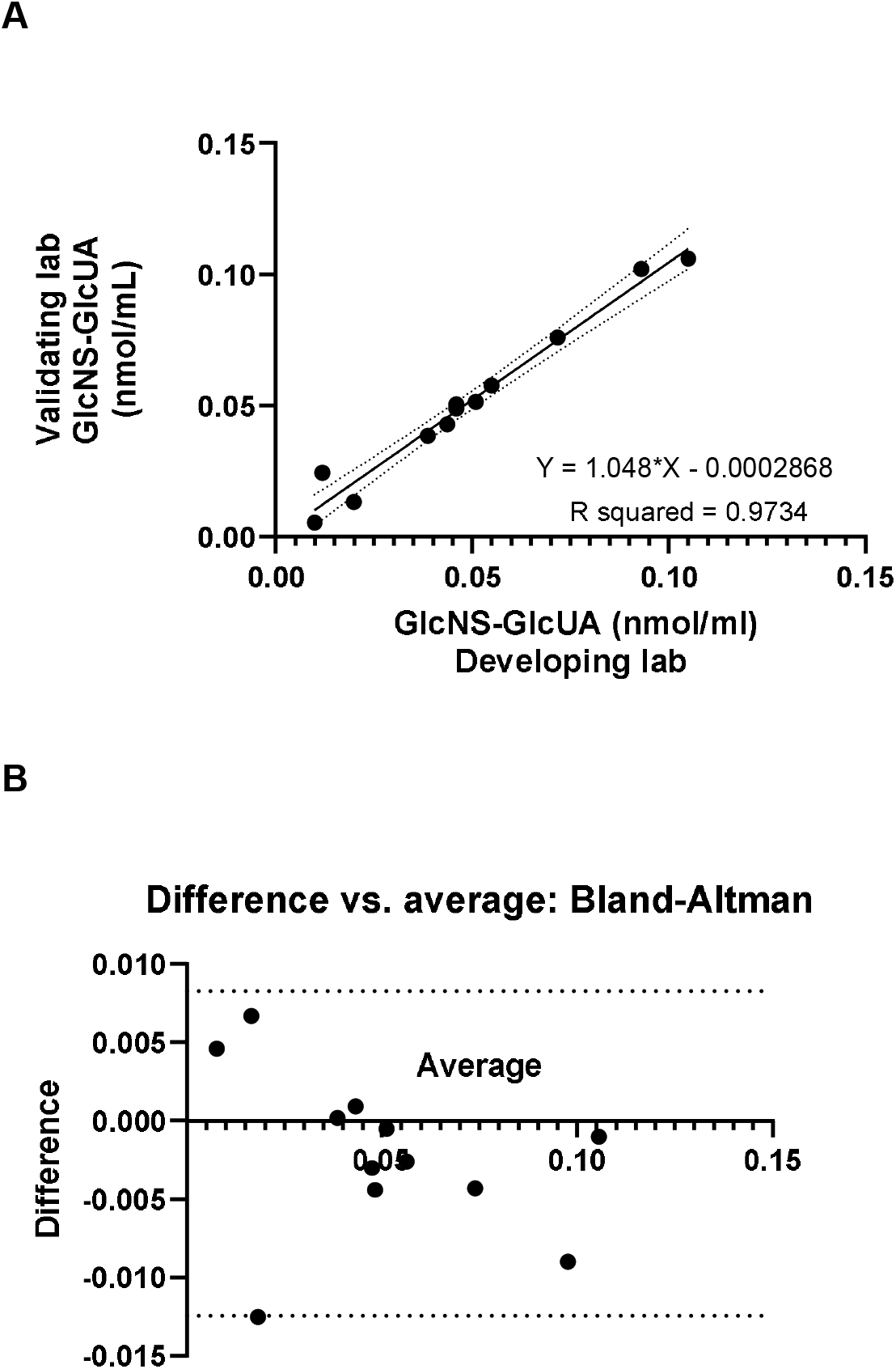
Intra-laboratory comparison between Ultragenyx (Development) and Veloxity Labs (Validation) using linear regression (A) and Bland-Altman comparison (B).

## 4. Conclusions

We developed and validated a quantitative LC-MS/MS assay for the HS-derived disaccharide GlcNS−GlcUA in human CSF. The method leverages a chemically defined reference standard and an isotopically labeled internal standard, PMP derivatization, PFP chromatography, and targeted MRM detection to deliver robust performance from 0.005 to 0.500 nmol/mL. Collectively, these data establish a regulatory-grade platform for quantitative CSF HS biomarker measurement, with evidence of good transferability to enable multiple labs to support clinical development.

The evolution in methodology and performance of GlcNS−GlcUA measurement provides a path to assay harmonization, enabling improved assessment of pharmacodynamic response to therapy. Because no treatment for MPS IIIA is currently available and patient CSF samples are limited, maximizing the ability of investigators to objectively compare CNS substrate burden across patients will enable optimal evaluation and deployment of emerging therapies for MPS IIIA.

## Data Availability

All data produced in the present study are available upon reasonable request to the authors.

## Acknowledgements

The authors acknowledge Henry Law (Ultragenyx Pharmaceutical Inc.) and Veloxity Labs for their contributions. Ben Scott, PhD (Scott Medical Communications, LLC) provided medical editing support funded by Ultragenyx Pharmaceutical Inc.

